# Detection of Early HIV-1 Infection among Febrile Persons and Blood Donors in Oyo State, Nigeria

**DOI:** 10.1101/2020.04.09.20058966

**Authors:** Babatunde A. Olusola, David O. Olaleye, Georgina N. Odaibo

## Abstract

About 37.9 million persons are infected with HIV globally resulting in 770,000 deaths. Over 50% of this infection and deaths occur in Sub-Saharan Africa with countries like Nigeria greatly affected. The country also has one of the highest rate of new infections globally. Diverse HIV-1 subtypes have been identified in the country. Febrile persons and blood donors pose a great transmission risk in the country especially during the early stages of infection. HIV-1 rapid kits are routinely used for diagnosis among the general population and high risk groups. However, there is limited information on the usefulness of HIV rapid kits for early detection especially in areas where diverse HIV-1 subtypes circulate. In this study, the prevalence of early HIV-1 infection as well as circulating HIV-1 subtypes among febrile persons and blood donors were determined. Furthermore, the sensitivity of a widely used HIV-1 rapid antibody kit was compared with those of Antigen/Antibody ELISA based methods. Participants were recruited from selected hospitals in Ibadan and Saki, Nigeria. The prevalence of early HIV infection among 1028 febrile persons (Ibadan: 2.22%; Saki: 1.36%) and blood donors (5.07%) studied were significantly different (P<0.03674). CRF02_AG was the predominant subtype detected with more diverse HIV-1 subtypes observed among febrile persons compared to blood donors. About 1.2% of the samples detected on Antibody based ELISA methods were undetectable on the HIV-1 rapid antibody kit. Genetic diversity of HIV-1 strains among infected individuals in Oyo State, Nigeria is still relatively high. This diversity is likely impacting on diagnosis.

## Introduction

As at 2018, about 37.9 million persons are infected with HIV globally with 21% of these people not aware of their status^1^. This rate of infection resulted in 770,000 deaths. Also, in the same year, 1.7 million (4.4%) individuals became newly infected with the virus. Over 50% of HIV infection and AIDS-related deaths occur in Sub-Saharan Africa making the region the highest hit with the infection^1^. Apart from several HIV-1 subtypes and recombinant forms circulating in Africa, more than a few high risk groups are also present in the continent^1,2^. Although HIV prevalence is controlled in the general population in most countries in Africa, there is still high prevalence of the virus among high risk groups. To control the HIV epidemic and achieve the 95-95-95 target, it is important that new HIV infections are quickly diagnosed and high risk groups are continuously identified and monitored^3^. Furthermore, it is pertinent that circulating subtypes within these groups are determined. Previous studies have showed that HIV-1 subtypes have differing impacts on diagnosis, transmission, pathogenesis and vaccine designs^2,4,5^.

In Nigeria, over 1.9 million people are infected with HIV and around 33% of these people don’t know their status. Although there is reduction in the prevalence of HIV infection, the country still has one of the highest number of new HIV infections (130,000) and AIDS-related deaths (53,000) globally^1^. Diverse HIV-1 subtypes and recombinant forms (CRF02_AG, G, A, B, C, D, CRF06_cpx, URFs etc.) have been characterized in the country previously^6^. Several HIV-1 high risk groups such as commercial sex workers^7,8^, voluntary blood donors^9,10^, long distance drivers^11–13^, mammy markets contiguous to military facilities^14^ etc. have also been identified. We as well as other authors have previously identified persons with febrile symptoms as a group to be screened for early HIV infection^15–17^. Most of these individuals are unaware of their status, hence, they pose an even higher transmission risk. The same applies to voluntary blood donors. Most blood screening centers in the country use HIV-1 Rapid antibody kits for pre-donation diagnosis. These kits miss out on infected persons at the early stages of infection. In order to sustain the gains of HIV control in the country, there is the need to diagnose persons at the early stages of HIV-1 infection as well as monitor high risk groups such as voluntary blood donors. Voluntary blood donors may fuel the spread of the virus as there is still high prevalence of unscreened or improperly screened HIV infected blood in Nigeria and other developing countries^18,19^.

Despite concerted efforts in the country to control HIV infection, an appreciable number of persons do not know their status and there is very limited data on circulating HIV-1 subtypes among febrile persons and voluntary blood donors. Although opportunities for HIV-1 testing have improved in the country, most testing centers use HIV-1 Rapid Antibody kits for diagnosis. These kits increase access by reducing turnaround time. However, their usefulness in eliminating samples with HIV-1 antibodies during early HIV infection diagnosis is not known. It is uncertain whether these kits’ sensitivities are not reduced in areas with high HIV-1 prevalence and diverse HIV-1 subtypes. Therefore, the aim of this study was to determine the prevalence and circulating subtypes of early HIV-1 infection among voluntary blood donors and febrile persons referred by clinicians for malaria antigen testing in hospitals in Ibadan and Saki, Oyo State, Nigeria. The performance characteristics of an HIV Rapid Antibody Point of Care test kit (Alere™ Determine™ HIV-1/2 set) for HIV-1 antibody detection was also determined.

## Materials and Methods

### Study sites and patient population

This study was conducted in two major cities in Oyo state, namely Ibadan and Saki as previously described^15^. Ibadan is the capital city of Oyo State and one of the most populated cities in Africa while Saki is a town 100 miles west of Ibadan, toward the border with the republic of Benin. Multiple HIV-1 subtypes and circulating recombinant forms (CRFs) have been shown previously to be circulating in these two cities^20^. Government, missionary and private owned hospitals in these cities were selected based on accessibility and availability of resident Physicians and laboratory staff. The Government hospitals were primary and secondary health care centers and services at these hospitals for outpatients were provided almost at little or no costs. Voluntary blood donors as well as persons referred by clinicians for malaria antigen test from selected hospitals in Ibadan and Saki were recruited for the study.

### Recruitment of participants, sample collection and processing

Participants were recruited for this study after obtaining informed consent. Data on patient age, gender and contact details were collected. They were given feedback on their results within a week of sample collection. Individuals infected with HIV were counseled and integrated into existing HIV point of care centers. We screened 1028 participants for this study out of which 890 were referred by Physicians to on-site laboratories for malaria antigen test. This study was a continuation of an initial study started in 2015. Initial findings from the study have been previously reported^15^. Five milliliters of whole blood were collected in EDTA bottles from participants. Plasma was separated from the samples immediately after collection, stored at −20°C and transported in cold chain to a central laboratory for analysis. The samples were then stored at −80°C until analyzed. Blood samples were analyzed for HIV antigen/antibody and HIV-1 Gag DNA.

### Identification of early and chronic HIV-1 infections

The updated CDC algorithm of laboratory testing for the diagnosis of early and chronic HIV-1 infection was used for this study as previously described^15^. Briefly, samples were identified as early HIV infection if HIV-1 DNA was detected after being positive with a 4th generation HIV ELISA (detection of both antigen and antibody) and negative with 3^rd^ generation HIV ELISA (detection of HIV antibody only) while those from which HIV-1 DNA scored positive with both 4^th^ and 3^rd^ generation ELISA, were classified as chronic HIV infection. Genscreen™ ULTRA HIV-Ag-Ab ELISA kit (Biorad, Hercules, California) was used as the 4^th^ generation ELISA kit while AiD™ anti-HIV 1+2 ELISA kit (Wantai, Beijing, China) was used as the 3^rd^ generation ELISA kit. All assays were performed under strict biosafety conditions and according to the manufacturer’s recommendation. Furthermore, HIV-1 antibodies was also determined using Alere™ Determine™ HIV-1/2 Rapid Antibody Test kit (HIV Rapid Ab POC) (Alere, Chiba, Japan) in a subset of the samples. This test kit is approved for routine HIV diagnosis in the country.

### PCR amplification and sequencing

Total DNA was extracted from whole blood using guanidium thiocyanate in house protocol. A fragment of the gag-pol region (900 base pairs) of the virus was amplified using previously published primers and cycling conditions ^21^ with slight modifications. Briefly, PCR was performed using a SuperScript III One-step RT-PCR system with platinum TaqDNA High fidelity polymerase (Jena Bioscience). Each 25µl reaction mixture contained 12.5µl reaction mix (2x), 4.5µl RNase-free water, 1µl each of each primer (20pmol/µl), 1µl Superscript III RT/Platinum Taq High Fidelity mix, and 5µl of template DNA. Pan-HIV-1_1R (CCT CCA ATT CCY CCT ATC ATT TT) and Pan-HIV-1_2F (GGG AAG TGA YAT AGC WGG AAC) were used. Cycling conditions were 94°C for 5min; 35 cycles of 94°C for 15s, 58°C for 30s, and 68°C for 1min 30s; and finally, 68°C for 10mins.

Positive HIV samples that was undetectable using the above stated primers were retested using another set of GAG primers for nested PCR as described previously^22^. Briefly, a 700-bp fragment corresponding to the p24 region was amplified using G00 and G01 for first round and G60-G25 for the second round. The PCR conditions were as follows: an initial denaturation step for 3 min at 92°C, followed by 30 cycles of 92°C for 10 s, 55°C for 30 s, and 1 min at 72°C, with a final extension for 7 min at 72°C. Amplicons were detected by gel electrophoresis. Furthermore, HIV-1 VIF gene of some samples were also detected and sequenced. As described previously^23^, Vif-1 (ATTCAAAATTTTCGGGTTTATTACAG) and TATED3-1 (AATTCTGCAACAACTGCTGTTTAT) primers were used for first round while Vif-2 (AGGTGAAGGGGCAGTAGTAATACA) and TATED-1(GCAGGAGTGGAAGCCATAATAAG) primers used for second round. The PCR conditions were as follows: 94°C for 5mins, followed by 35 cycles of 94°C for 30s, 60°C for 30s, 68°C for I min with a final extension for 4min at 68°C for 1minPositive PCR reactions were shipped on ice to Macrogen, South Korea for Big Dye sequencing using the same amplification primers (Pan-HIV-1_1R and Pan-HIV-1_2F; or G60 and G25; or Vif-2 and TATED-1 for VIF).

### Detection of HIV-1 subtypes and recombination

The sequences were cleaned and edited using Chromas and Bioedit softwares. Subtyping was performed using a combination of four subtyping tools: The Rega HIV-1 Subtyping Tool, version 3.0 (http://dbpartners.stanford.edu/RegaSubtyping/), Comet, version 2.2 (http://comet.retrovirology.lu), National Center for Biotechnology Information, Bethesda, MD (http://www.ncbi.nlm.nih.gov/Blast.cgi) and jpHMMM: Improving the reliability of recombination prediction in HIV-1 (http://jphmm.gobics.de/submission_hiv). The first three tools were used simultaneously while jpHMMM was used to resolve discordant subtypes. Phylogenetic analyses were performed using MEGA software version 10. All consensus nucleotide sequences obtained in this study were submitted to GenBank. Database and assigned accession numbers KY786266-272 and MN943617-635.

### Ethical approval

Ethical approvals for this research were obtained from the University of Ibadan/ University College Hospital (UI/UCH) Research and Ethics Committee (UI/EC/15/0076) and the Oyo State Ministry of Health Committee on Human Research (AD13/479/951). All results were delinked from patient identifiers and anonymized.

### Eligibility/ Exclusion criteria

Only individuals between 18 and 65 years of age were included in the study. Individuals who already knew their HIV status were excluded from the study.

### Data management and statistical analysis

Statistical analyses were performed using SPSS version 20 and graphpad prism. Categorical variables were compared using Z and chi-square tests while quantitative variables were compared using ANOVA. Test of significance was set at P<0.05.

## Results

### Patient characteristics

One thousand and twenty-eight (1028) individuals participated in this study out of which 890 were outpatients referred for malaria antigen testing. Voluntary blood donors were only recruited in Ibadan. Five hundred and ninety four (57.7%) of these individuals were female. The mean (±SD) ages for participants referred for malaria antigen test in Ibadan, Saki and voluntary blood donors were 37.2 (±9.1), 36.1 (±14.3) and 35.1 (±14.3) years respectively (Table 1).

**Table 1:**
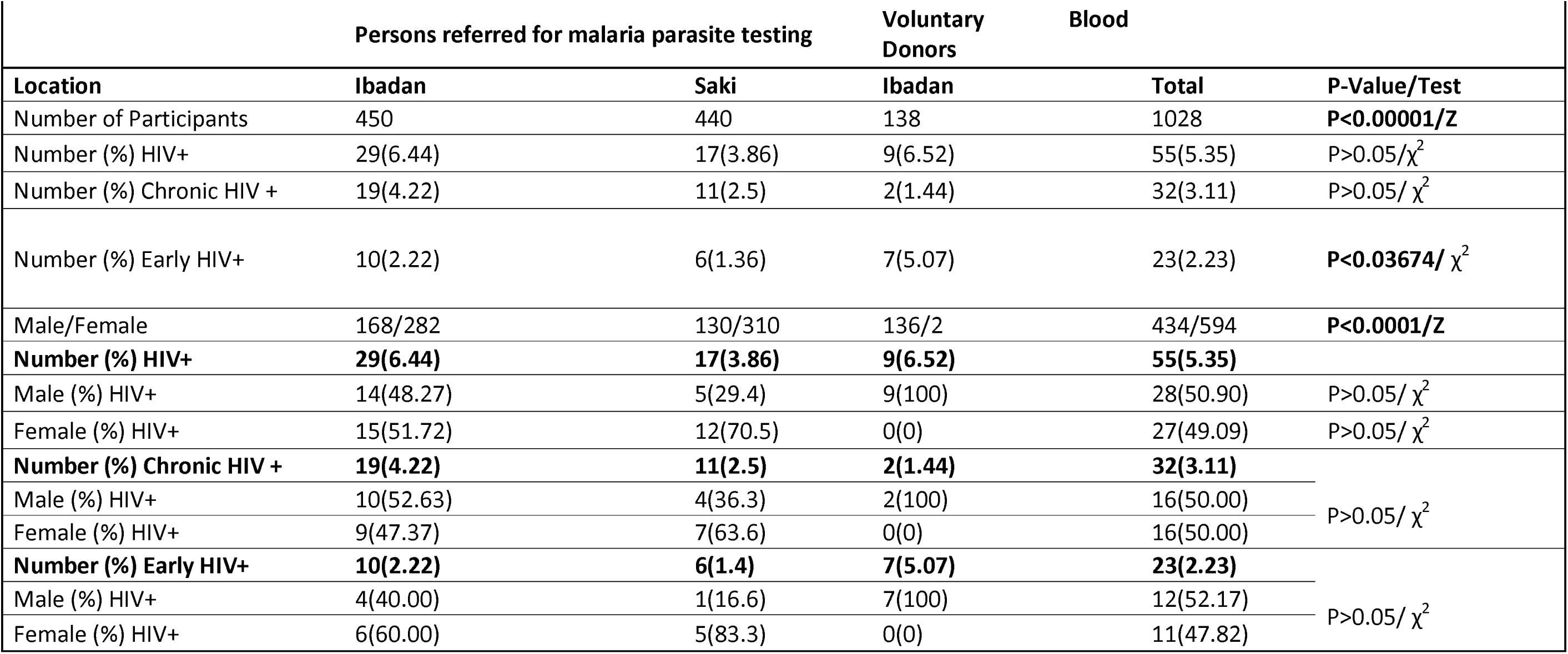
Prevalence of early and chronic HIV-1 infection among Persons referred from malaria parasite testing and Voluntary Blood Donors. Values shown are in number and percentages. Statistical tests used: Z and chi-square (χ^2^) tests. Significant differences are in bold fonts while differences that are not significant are written as P>0.05.

### Prevalence of early and chronic HIV infection

A total of 55 persons were HIV positive, giving a rate of infection of 5.35%. The HIV prevalence varied by location and groups tested. Twenty three (2.23%) individuals were at the early stages of HIV infection with an higher rate among voluntary blood donors (5.07%) compared to those referred for malaria antigen test (Ibadan: 2.22%; Saki: 1.36%) (P<0.03674). Differences in prevalence by gender were not significant for either early or chronic HIV infection (Table 1).

### Proportions of HIV reactive samples based on serological assays

Table 2 shows the proportions of HIV reactive sample based on serological assays. This analysis was carried out on a subset of samples (671) collected from persons referred for malaria antigen test in Ibadan and Saki. Using 4^th^ generation HIV Ag-Ab ELISA as the gold standard, HIV Rapid Ab POC (Ibadan: 1.95%; Saki: 2.95%) had significantly higher rates of false negative results in both locations compared to 3^rd^ generation ELISA (HIV Ab ELISA) (Ibadan: 1.29%; Saki: 1.36%). The performance characteristics of HIV Rapid Ab POC are also presented. The sensitivity (95% CI) of HIV Rapid Ab POC is 52.38% (29.78-74.29). As shown in Figure 1, 14 samples were reactive on all serological assays while no sample was reactive on HIV Ab ELISA alone nor on HIV Ab ELISA and HIV Rapid Ab POC. Eight samples with HIV-1 antibodies (1.2%) were undetectable on HIV Rapid Ab POC.

**Table 2:** Proportions of HIV reactive samples based on Enzyme Immunoassay. Values shown are in number and percentages. *Difference in numbers of reactive HIV samples between Alere™ Determine™ HIV-1/2 Rapid Antibody Test kit (HIV Rapid Ab POC) and 4^th^ generation ELISA kit (HIV Ag/Ab ELISA) for samples collected in Ibadan was significant (P<0.0001). ^#^Difference in numbers of reactive HIV samples between HIV Rapid Ab POC and HIV Ag/Ab ELISA for samples collected in Saki was significant (P<0.0001). ^a^Difference in numbers of reactive samples (using HIV Ag/Ab ELISA) between Ibadan and Saki was significant (P<0.001). χ^2^ was used to test for significance. a= True Positive; b= False Negative; c= False Positive; d=True Negative.

| Location | Number Tested | HIV Rapid Ab POC |  | HIV Ab ELISA |  | HIV Ag/Ab ELISA |
| --- | --- | --- | --- | --- | --- | --- |
|  |  | Reactive (%) | False Negative (%) | Reactive (%) | False Negative (%) | Reactive (%) |
| Ibadan | 231 | 8(3.46)* | 6(1.95) | 11(4.76) | 3(1.29) | 14(6.06)* |
| Saki | 440 | 8(1.81)<sup>#</sup> | 9(2.95) | 11(2.50) | 6(1.36) | 17(3.86)<sup>#</sup> |
| Total | 671 | 16(2.38) | 15(2.45) | 22(3.27) | 9(1.34) | 31(4.61)<sup>a</sup> |
|  |  |  |  |  |  | *P<0.0001 |
|  |  |  |  |  |  | <sup>#</sup>P<0.0001 |
|  |  |  |  |  |  | <sup>a</sup>P<0.001 |
| **HIV Rapid Antibody** |  |  |  |  |  |  |
| Sensitivity (95% CI) | 52.38% (29.78-74.29) | a/a+b |  |  |  |  |
| Specificity (95%CI) | 99.50%(98.20-99.94) | d/c+d |  |  |  |  |
| PPV (95%CI) | 84.62%(54.55-98.08) | a/a+c |  |  |  |  |
| NPV (95%CI) | 97.54%(95.53-98.82) | d/b+d |  |  |  |  |

**Figure 1:**
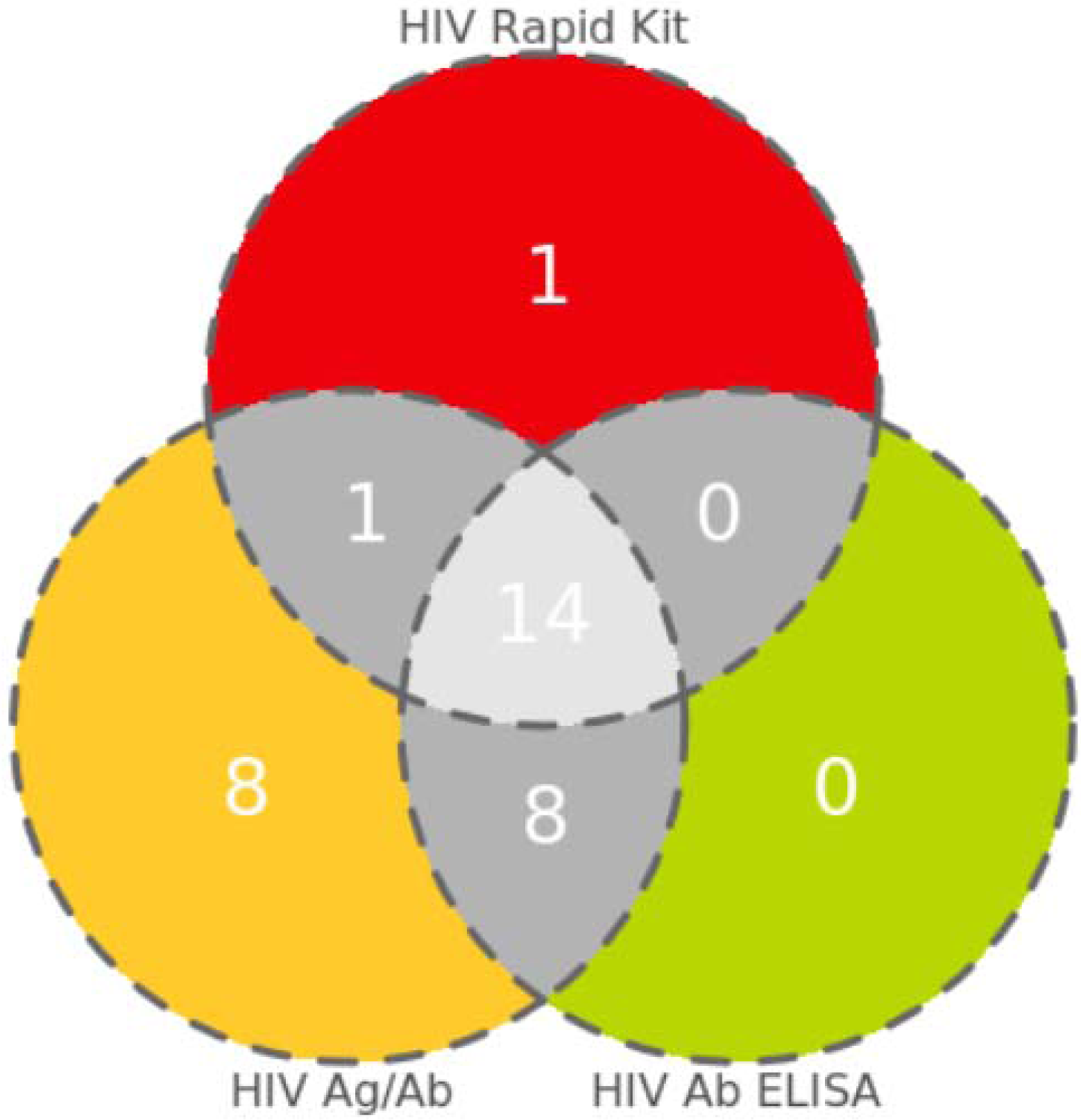
Diagram showing number of HIV positive samples on serological assays. Values are shown in numbers only. Total number of samples positive for an assay type corresponds to the addition of all numbers in the circle representing the assay. Samples positive on Alere™ Determine™ HIV-1/2 Rapid Antibody (HIV Rapid Kit), 3^rd^ generation ELISA kit (HIV Ab ELISA) and 4^th^ generation ELISA kit (HIV Ag/Ab) are represented in red, green and yellow colours respectively. Those positive on two assays are in grey while samples positive on the three assays are in light grey. No sample was reactive on 3^rd^ generation HIV ELISA (HIV Ab ELISA) alone. Also no sample was reactive on both Alere™ Determine™ HIV-1/2 Rapid Antibody (HIV Rapid Kit) and HIV Ab ELISA.

**Figure 2:**
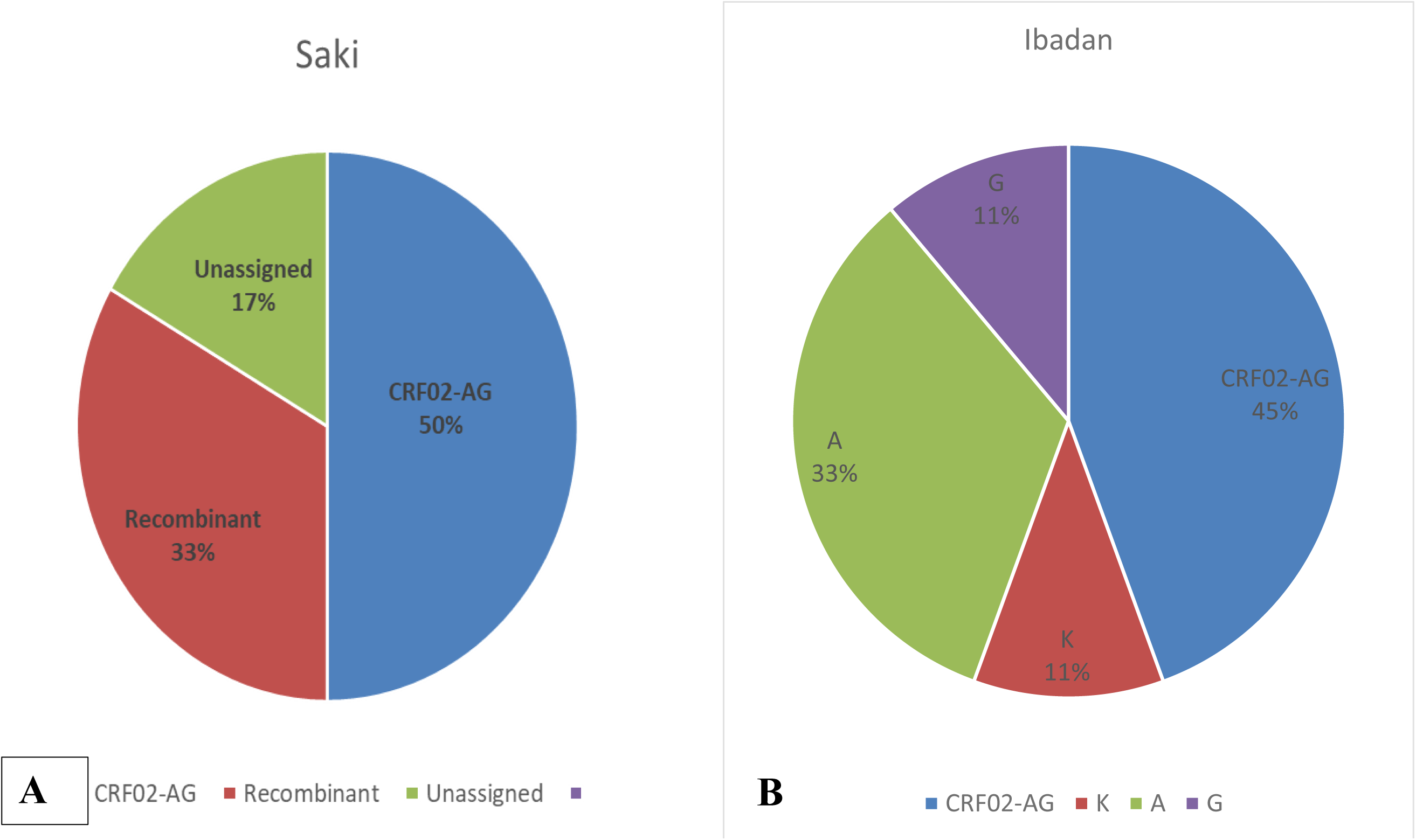
Summary of subtypes isolated from individuals at the early stages of HIV-1 infection. Values are shown in percentages only. **A**. Pie chart showing the distribution of subtypes isolated from persons referred for malaria parasite antigen testing in Saki, Nigeria. Total number of individuals at this location is 6. One sample was not sequenced. Three, 1 and 1 samples were subtyped as CRF-02AG, Recombinant GD and Unassigned respectively. **B**. Pie chart showing the distribution of subtypes isolated from persons referred for malaria parasite antigen testing in Ibadan, Nigeria. Total number of individuals at this location is 10. One sample was not sequenced. Four and 3 samples were subtyped as CRF-02AG and A respectively while one sample were subtyped as G and K each.

**Figure 2C:**
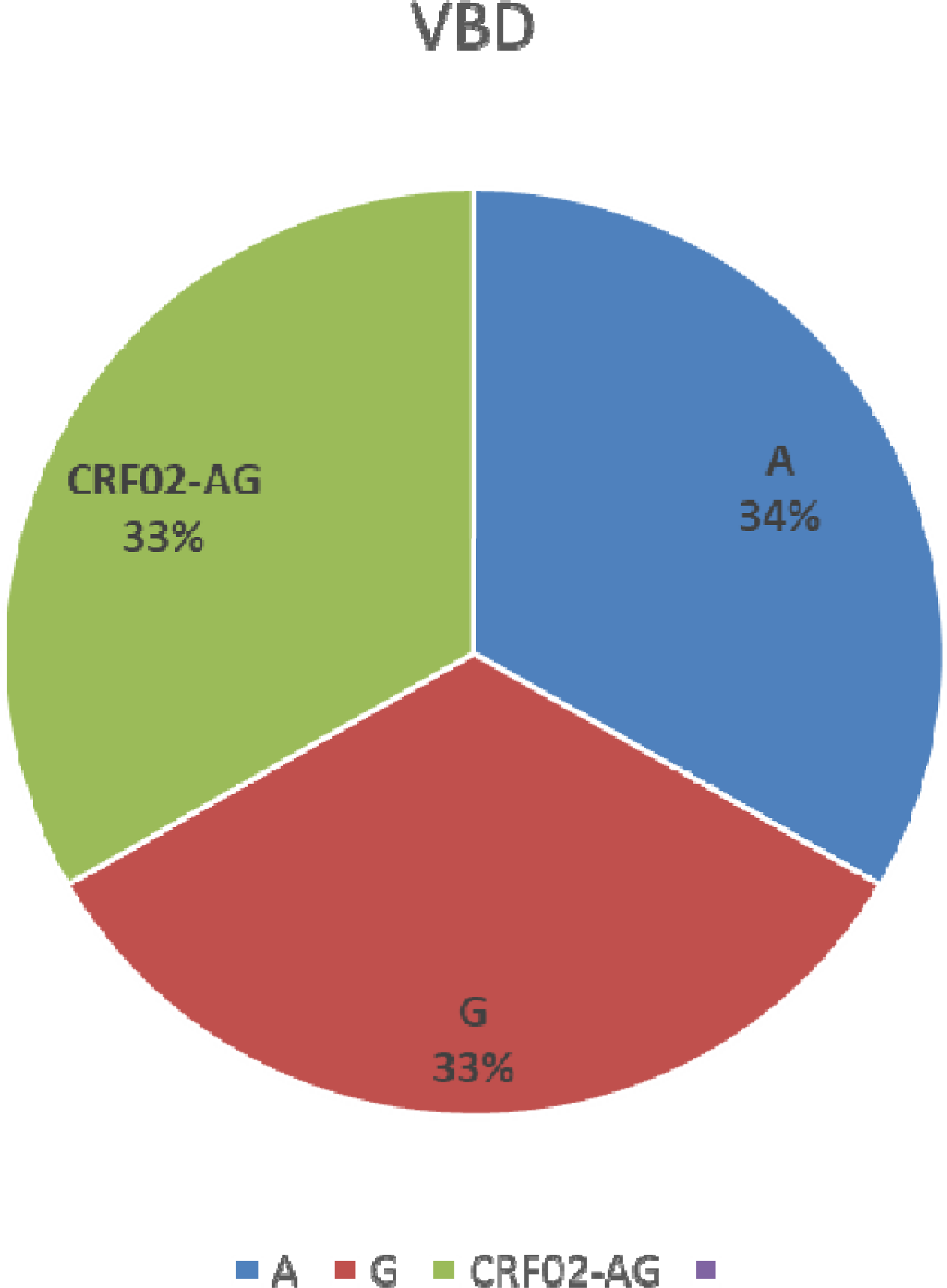
Summary of subtypes isolated from individuals at the early stages of HIV-1 infection. **C**. Pie chart showing the distribution of subtypes isolated from Voluntary blood donors in Ibadan, Nigeria. Total number of individuals in this group is 7. One sample was not sequenced. Two samples each were subtyped as A, G and CRF02-AG.

### Subtyping

Twenty out of the 23 isolates with early HIV infection were successfully sequenced. The predominant subtype detected was CRF02_AG (9; 45%). Other subtypes detected were A (5; 25%), G (3; 15%), K (1; 5%) and Recombinant GD (1; 5%). One of the sequenced viruses did not align with any subtype and was reported as unassigned (Table 3). Based on groups, CRF02-AG was the most predominant strain among persons referred for malaria antigen test (Saki: 50%; Ibadan 40%). Voluntary blood donors had the same number of HIV-1 subtypes A, G as well as CRF02-AG. Subtype K, recombinants and unassigned subtypes were found among persons referred for malaria antigen test.

**Table 3:** Subtype classification of HIV-1 GAG gene isolated from individuals at the early stage of infection. Letter a in superscript corresponds to samples that the GAG gene were not sequenced, rather VIF gene was sequenced and used for subtyping.

| Location | Sequence ID | Sub-typing tool |  | Accession number | Assigned subtype |
| --- | --- | --- | --- | --- | --- |
|  |  | REGA | COMET |  |  |
| Saki | EHIV001 | Recombinant | CPZ | KY786266 | Unassigned |
| Ibadan | EHIV002 | Recombinant of FK | CPZ | KY786267 | K |
| Saki | EHIV003 | CRF02-AG | CRF02-AG | KY786268 | CRF 02-AG |
| Saki | EHIV004 | CRF02-AG | A1/Check for 02-AG | KY786269 | CRF 02-AG |
| Saki | EHIV005 | CRF02-AG | CRF02-AG | KY786270 | CRF 02-AG |
| Saki | EHIV006 | Recombinant of GD | CPZ | KY786271 | Recombinant GD |
| Ibadan | EHIV007 | CRF02-AG | CRF02-AG | KY786272 | CRF 02-AG |
| Saki | EHIV008 |  |  | Not Sequenced |  |
| Ibadan | EHIV009 |  |  | Not Sequenced |  |
| Ibadan | EHIV010 | A1 | A1 | MN943617 | A |
| Ibadan | EHIV011 | A1 | A1 | MN943616 | A |
| Ibadan | EHIV012 | G | G | MN943624 | G |
| Ibadan | EHIV013 | A1 | A1 | MN943613 | A |
| Ibadan | EHIV014 | CRF02-AG | CRF02-AG | MN943628 | CRF 02-AG |
| Ibadan | EHIV015 | A1 | A1 | MN943615 | A |
| Ibadan | EHIV016 | G | G | MN943627 | G |
| Ibadan | EHIV017 | CRF02-AG | CRF02-AG | MN943629 | CRF 02-AG |
| Ibadan | EHIV018 | CRF02-AGvif | CRF02-AGvif | MN943633a | CRF 02-AG |
| Ibadan | EHIV019 |  |  | Not Sequenced |  |
| Ibadan | EHIV020 | CRF02-AGvif | CRF02-AGvif | MN943634a | CRF 02-AG |
| Ibadan | EHIV021 | CRF02-AGvif | CRF02-AGvif | MN943635a | CRF 02-AG |
| Ibadan | EHIV022 | G | G | MN943625 | G |
| Ibadan | EHIV023 | A1 | A1 | MN943619 | A |

<sup>a</sup>Gag gene was not sequenced for these samples

## Discussion

This study indicates that the prevalence of early HIV-1 infection is significantly high among febrile persons referred for malaria antigen testing (Ibadan: 2.22%; Saki: 1.36%) and voluntary blood donors (5.07%). Presently, the prevalence of HIV-1 among the general population in Nigeria is about 1.9%^24^. This then means that high prevalence of HIV-1 infection among high risk groups is masked by low prevalence in the general population as previously hypothesized^25^. To effectively control the prevalence and incidence of HIV-1 infection in the country, it is important that persons at the early stages of infection are identified and placed on treatment early. The strategy used in this study to identify early HIV-1 infection seems applicable for other high risk groups’ aside febrile persons. Identifying and treating HIV-1 infected individuals during the early stages of infection will go a long way in reducing new HIV-1 infections as well as decrease the number AIDS-related deaths in the country, since undetectable equals untransmittable^26^. Furthermore, results of this study show that CRF02-AG, subtypes A and G in that order still remains the predominant circulating HIV-1 subtypes in Oyo state as well as among high risk groups in the state. These strains have been shown to be circulating in the region since 1995^20,27^. We also showed that HIV-1 Rapid antibody kit is not advisable for use as a diagnostic tool for early HIV-1 infection. A previous study in Nigeria that characterized acute HIV-1 infection among high risk groups had a lower prevalence (0.05%) compared to what was observed in this study^28^. This may be due to the fact that HIV-1 Rapid Antibody was used to eliminate seroconversion in the study.

Results of HIV-1 prevalence in this study is very similar to what was obtained from our previous study among persons referred for malaria antigen test^15^. Furthermore, our results on the prevalence of HIV-1 infection among blood donors is not different from previous studies on this group that used ELISA based tests for diagnosis^9,19,29^. Previous studies on HIV-1 prevalence among blood donors in Nigeria have reported varying proportions^9,30–38^. Aside the fact that various regions in the country have varying HIV-1 prevalence^24^, the method used for HIV-1 diagnosis played a significant role in the numbers of infected persons detected. In 2008, Odaibo^39^ *et al*., reported that 6(1.2%) out of 500 antibody-negative blood donor samples had HIV-1 antigens and cDNA. This report is not different from what was observed in this study in which 8(1.2%) samples positive on HIVAg/Ab ELISA were undetectable on Antibody based testing kits (see Figure 1). The case is really worsened for HIV-1 Rapid POC used in this study in which 16 (2.4%) samples positive on either HIV Ag/Ab ELISA and/or HIV Ab ELISA were undetectable on the platform. The average score of HIV Rapid POC sensitivity observed in this study is similar to what was observed by Mba *et al*., 2018^40^. In the study which was carried out among blood donors in Gabon, the authors argued that Rapid antibody kits may not be very useful for pre-donation HIV diagnosis.

It is important that blood donation testing centers use HIV Antigen and Antibody based ELISA tests for pre donation screening at the minimum. Other authors have showed the cost benefits of using only HIV antigen and antibody based ELISA test kits for pre donation blood screening rather than the two step algorithm involving the use of HIV Rapid antibody kits^41^. Findings from this study suggest that missed opportunities for HIV-1 diagnosis still occur even after the presence of detectable antibodies in blood samples. Reasons for this may be that serological methods for detecting HIV-1 infection have different sensitivities across various circulating subtypes. Although we could not associate specific sensitivity scores to HIV-1 screening methods against the various subtypes identified in this study, we have showed that HIV-1 Rapid antibody kits may not be very effective in detecting HIV-1 antibodies in areas where multiple HIV-1 subtypes circulate. This missed diagnosis leads to further transmission of the virus to susceptible individuals. It is unadvisable to use HIV-1 Rapid antibody kits for pre-donation screening in regions with diverse circulating HIV-1 subtypes^40^.

We have previously reported the detection of CRF02_AG, subytpe K, recombinant strain GD and an unassigned subtype among persons referred for malaria parasite testing (KY786266-272)^15^. In this study, more CRF02_AG, subtypes A and G were identified among febrile persons and blood donors. As earlier stated, CRF02_AG remains the predominant subtype among high risk groups in Oyo State. The strain is the predominant circulating virus in West Africa^6,20,27,42,43^. Although CRF02_AG was first described in Ibadan, Nigeria, the origin of the virus is still not known especially now that the strain is prevalent in several regions of the world. Subtypes A and G have also been previously reported as circulating in Oyo state as well as in other parts of Nigeria. A recent study identified CRF02_AG, as well as subtypes G and A as the predominant circulating HIV-1 strains in Ibadan, Nigeria^6^.

It is important that in depth analysis of HIV-1 viral diversities in geographical regions are carried out. This is because subtypes differences have been associated with varying patterns of replicative capacities, diagnosis, transmission and pathogenesis. In another study, we have showed that HIV-1 strains circulating in Oyo State are associated with renal dysfunctions and reduced proportions of T cells in infected individuals during early stages of HIV-1 infection^44^. Findings in this study observed more subtypes in Ibadan metropolis (urban settlement) compared to Saki town (rural settlement). Recent studies using phylodynamic tools have suggested that HIV-1 strains originate and expand in urban areas before migrating to smaller rural centers^6^. There is the need to describe the phylodynamics of HIV-1 subtypes in other states of the country as this will shed further light into the diversity of the virus.

In summary, we have showed that the genetic diversity of HIV-1 strains among infected individuals in Oyo State, Nigeria is still relatively high. This diversity is likely impacting on diagnosis. Our study also suggests that screening persons for early HIV-1 infection is best done using Antigen/Antibody based ELISA techniques for elimination of seroconversion. Early HIV-1 infected individuals are the major drivers of new infections. Hence, the need to use highly sensitive diagnostic tools for HIV-1 identification among them. This is particularly important for high risk groups such as voluntary blood donors. Studies examining the impact of host immune pressures on the generation of HIV-1 immune escape mutants during the early stages of infection are ongoing.

## Data Availability

Data will be made available by the corresponding author upon request.

## Acknowledgements

We thank all individuals who participated in this study. Special appreciation to Mrs. Fadimu and Mrs. Adeyefa (both of Blood bank, University College Hospital, Ibadan, Nigeria) for linking us up with volunteer blood donors. This study was supported by University of Ibadan MEPI Junior Faculty Research Training Program (UI-MEPI-J) mentored research award through National Institute of Health (NIH) USA grant funded by Fogarty International Centre, the office of AIDS Research and National Human Genome Research Institute of NIH, the Health Resources and Services Administration (HRSA) and the Office of the U.S. Global AIDS Coordinator under award number D43TW010140 to BO. The funders had no role in study design, data collection and analysis, decision to publish, or preparation of the manuscript. The authors have no conflict of interest to declare.

## Sequence Notes

All consensus nucleotide sequences obtained in this study were submitted to GenBank. Database and assigned accession numbers KY786266-272 and MN943617-635.

## Reprints requests

Reprints requested should be directed to Prof Gerogina N. Odaibo, Department of Virology, College of Medicine, University College Hospital, Ibadan, Nigeria. foreodabo@hotmail,com

## Notes

### Competing Interest Statement

The authors have declared no competing interest.

